# The effect of self-management online modules plus nurse-led support on pain and quality of life among young adults with irritable bowel syndrome: A randomized controlled trial

**DOI:** 10.1101/2022.02.23.22271431

**Authors:** Jie Chen, Yiming Zhang, Zahra Amirkhanzadeh Barandouzi, Joochul Lee, Tingting Zhao, Wanli Xu, Ming-Hui Chen, Bin Feng, Angela Starkweather, Xiaomei Cong

**Author notes:** Corresponding author: Xiaomei Cong, PhD, RN, FAAN, Professor and Associate Dean for Research Director, Biobehavioral Research Laboratory University of Connecticut School of Nursing 231 Glenbrook Road, Unit 4026, Storrs, CT 06269-4026.

## Abstract

**Background:** Irritable bowel syndrome (IBS) is a chronic pain condition that needs life-long self-management. However, the effect of self-management among young adults with IBS is limited.

**Objectives:** This study aimed to examine the effect of a nurse-led self-management program on IBS related pain and symptoms, and quality of life (QOL) among young adults with IBS.

**Theoretical framework:** The Individual and Family Self-Management Theory (IFSMT).

**Design:** A randomized controlled trial with data collected at baseline, 6- and 12-week follow up.

**Settings and participants:** Eighty young adults with IBS recruited from two campuses of a public university and two gastrointestinal clinics were randomly assigned into a Self-Management Online education and learning Modules group (SMOM, n = 41) or a Nurse-Led SMOM group (NL + SMOM, n = 39). Twenty-one healthy controls (HCs) were also recruited from these two campuses.

**Methods:** All the IBS participants received the SMOM after baseline data collection. Participants in the NL + SMOM received additional three nurse-led one-to-one consultations at baseline, 6- and 12-week follow up. Self-reported pain, symptoms, IBS-related QOL, self-efficacy for managing chronic disease, and coping were measured at baseline, and 6- and 12- week follow up among the IBS participants. The HCs completed data collection of pain and symptoms at baseline and 12-week follow up. The intervention effects across study time points and the comparisons between the two interventional groups were analyzed using linear mixed models. A longitudinal mediation analysis was also conducted to explore the mediation effects of self-management mechanisms of the interventions.

**Results:** Both the SMOM and NL + SMOM groups showed significant interventional effects on decreasing pain intensity and pain interference and increasing IBS-QOL among young adults with IBS at the 12-week follow up (all p < 0.05). The NL + SMOM also had significant effect on reducing anxiety and greater improvement in IBS-QOL compared with the SMOM at the 12- week follow up (both p < 0.05). Increased self-efficacy mediated the intervention effect of the NL + SMOM on reducing pain interference and improving IBS-QOL, while the effect of the SMOM was mediated through decreased an inefficient coping strategy-catastrophizing.

**Conclusions:** Guided by the IFSMT, this study showed that both the pain self-management online education and nurse-led interventions were effective for alleviating pain and improving QOL among young adults with IBS by targeting the self-management process. The nurse-led program had a better outcome than the online education alone in improving IBS-QOL. **Registration number**: <u>NCT03332537</u>

**What is already known about the topic:**

- Irritable bowel syndrome (IBS) is a chronic condition warranting lifelong self- management.
- Unrelieved abdominal pain is associated with increased healthcare expenditures and decreased quality of life (QOL) in young adults with IBS.
- Self-management interventions have moderate effect on attenuating IBS related pain and symptoms.

**What this paper adds:**

- Both the IBS Self-Management Online education and learning Modules (SMOM) and Nurse-Led SMOM (NL + SMOM) developed in this study were efficient in reducing pain intensity and pain interference and ameliorating IBS-QOL among young adults with IBS.
- The NL + SMOM had a greater interventional effect on improving IBS-QOL compared with the SMOM alone.
- The NL + SMOM had an indirect effect on pain and QOL by increasing self-efficacy, while the indirect effect of SMOM on pain and QOL was derived by decreasing inefficient coping (e.g., catastrophizing).

## Introduction

Irritable bowel syndrome (IBS) is one of the most prevalent disorders of the gut-brain interaction that affects up to 8-12% of the population worldwide (Ford et al., 2020; Lacy & Patel, 2017; Sperber et al., 2017). IBS occurs in women more than in men and is more commonly diagnosed in people younger than 50 years old (Farmer & Aziz, 2013). The economic impact of IBS is substantial and varies among different countries. The estimated costs range from $2 billion per annum in China, £45.6-200 million per annum in the UK, and between $1,562 and $7,547 per patient annually in the United States (Black & Ford, 2020). IBS is often characterized by recurrent abdominal pain, bloating, and altered bowel habits with diarrhea and/or constipation in the absence of demonstrable organic disease (Alammar & Stein, 2019). IBS symptoms can occur due to combination of different factors, including visceral hypersensitivity, altered bowel motility, neurotransmitters imbalance, infection, and psychosocial factors (Chen et al., 2022; Saha, 2014). IBS symptoms, while not life-threatening, post great burdens on both patients and society, the management of which is a global challenge for healthcare systems (Moayyedi et al., 2017). However, self-management interventions among young adults with IBS that incorporate standardized symptom measurements are limited and demanded (Cong, Perry, et al., 2018).

Recurrent abdominal (visceral) pain has been recognized as a cardinal symptom of IBS (Drossman et al., 2009; Page et al., 2018). Visceral pain is a generic term that describes pain originating from internal organs within the thorax and abdomen (Delvaux, 2002; Moloney et al., 2016). In normal conditions, nociceptors sense painful stimuli and project signals onto spinal nociceptive neurons, which gets relayed to the brain to evoke the perception of pain. The brain generates an efferent signal back to the periphery exerting either an inhibitory or a faciliatory effect on the pain sensation (Chang, 2005). In IBS, repeated or chronic activation of the nociceptors due to various factors such as inflammatory mediators (e.g., chemokines, cytokines, corticotropin-releasing hormone, serotonin histamine, proteases, prostaglandins), brain dysfunction and abnormal interaction of the gut-brain axis leads to both peripheral and central sensitization of the brain-gut axis and results in chronic visceral pain (Enck et al., 2016; Greenwood-Van Meerveld & Johnson, 2017; Widgerow & Kalaria, 2012).

Patients with IBS often relate the onset or aggravation of visceral pain to stress (Barbara et al., 2011). Stress can activate the mucosal mast cells of the gut and stimulate release of mediators such as serotonin and pro-inflammatory cytokines, which are responsible for the altered intestinal sensation and motility (Qin et al., 2014). Some evidence also suggests that psychological disturbances can contribute to IBS pain (Chen et al., 2022; Saha, 2014).

Supporting this, 70-90% of people with IBS report one or more psychiatric comorbidities such as anxiety and depression, which may exacerbate IBS symptoms (Barandouzi et al., 2022; Kopczyńska et al., 2018; Tosic-Golubovic et al., 2010). Moreover, IBS patients comorbid with depression experience low quality of life (QOL) (Kopczyńska et al., 2018). Given the importance of visceral pain in IBS and its extensive consequences, pain relief has been the main focus in IBS management (Chey et al., 2015; Ford et al., 2014).

Various approaches have been introduced for IBS-related pain and symptom management. Non-pharmacological interventions such as diet modifications, physical activity, and psychological therapy build on the therapeutic relationship between the patients and healthcare providers, which is an essential component of treatment recommended for the IBS management (Rawla et al., 2018). Patient-centered complementary and integrative medicine identifies a patient as the key player in pain management (Lee et al., 2014). In alignment, the concept of self-management places the patient as the central decision-maker in using self-regulation knowledge and skills as well as resulting actions for managing their health (Barlow et al., 2002). Self-management (SM) has been defined as *“a process by which individuals and families use knowledge and beliefs, self-regulation skills and abilities and social facilitation to achieve health-related outcomes”* (Ryan & Sawin, 2009). There are diverse formats of SM interventions that have been used in previous studies (Cong, Perry, et al., 2018). Internet-based interventions have been identified as one of the most efficient tools in SM for IBS (Pedersen, 2015). Studies also support other SM interventions such as self-training booklets, individual and group interventions, cognitive behavioral therapy (CBT), and other psychologic approaches as he fundamental features for building self-regulation knowledge and skills (Jarrett et al., 2016; Niesen et al., 2018; Shahabi et al., 2016). Nurse, as one of the most trusted healthcare provider, can help patients achieve better health outcomes by using self-regulation strategies based in SM theory (Grady & Gough, 2014). Evidence also supports that SM led by nurses helped individuals with IBS to effectively adapt and improve their QOL (Cong, Perry, et al., 2018; Pedersen, 2015).

The present study evaluated whether a nurse-led SM intervention can help individuals with IBS to improve their pain and symptom management as well as their QOL compared to only providing the self-Management Online education and learning Modules (SMOM). The Individual and Family Self-Management Theory (IFSMT) (Ryan & Sawin, 2009) was adopted as a theoretical framework of this study. Pain in young adults with IBS was considered as a component of the self-management context. The targeted interventions, developed from prior research and work with the patient population by us (Cong, Ramesh, et al., 2018), focused on the self-management process including self-efficacy and coping. IBS related symptoms were selected as proximal outcome variables, and pain and IBS related QOL were selected as distal outcome variables. The hypotheses included: 1) Both the self-Management Online education and learning Modules (SMOM) and the Nurse-Led one-to-one consultation plus the SMOM (NL + SMOM) interventions would decrease IBS related pain and symptoms, and improve quality of life (QOL) among young adults with IBS; 2) In comparison to the SMOM alone, the NL + SMOM intervention would have better effect on managing IBS related pain and symptoms, and on enhancing IBS-QOL; 3) The effects of the SMOM and NL + SMOM interventions on improving pain and symptom management and increasing QOL would be modified through increasing coping strategies and self-efficacy for managing chronic disease.

## Methods

### Design

A randomized controlled trial (RCT) was conducted to examine the effect of the SMOM alone versus the NL + SMOM intervention on pain, symptoms, and quality of life among young adults with IBS over a 12-week study period with data collected at baseline (T0), 6-week (T1), and 12-week (T2) follow-up visits. The study protocol was approved by the institutional review board (IRB) of a major research-intensive University in the northern Atlantic region (No. H16- 152). This RCT has been registered (<u>NCT03332537</u>) and the protocol was published (Cong, Ramesh, et al., 2018). A group of healthy young adults were also recruited in the study to serve as healthy controls (HCs), which received no intervention provided by the research team. The recruitment and data collection were conducted from October 2016 to March 2019.

### Settings and participants

IBS participants were recruited by posted flyers at gastrointestinal clinics in two hospitals and two campuses of a public university in the northeastern of United States. Healthy control (HC) subjects were recruited only from the college campuses and surrounding neighborhoods.

Volunteers were instructed to call a study-designated line for eligibility screening. All the data collections and nurse-led one-to-one consultations were allocated in a research laboratory affiliated with a university sponsored pain research center on the two campuses.

Young adults were eligible to enroll if they were: 1) aged 18 to 29 years older; 2) having IBS diagnosed by a healthcare provider based on Rome-III criteria; 3) able to access internet; 4) able to read and speak English; and 5) willing to participant in the study. Subjects were excluded if they had: 1) chronic pain conditions other than IBS including but not limited to chronic pelvic pain, or chronic interstitial cystitis; 2) infectious diseases (e.g., hepatitis, HIV, methicillin-resistant Staphylococcus aureus); 3) celiac disease or inflammatory bowel disease; 4) diabetes mellitus; 5) serious mental health conditions (e.g., bipolar disorder, schizophrenia, and mania); 6) regular use of opioids, iron supplements, prebiotics/probiotics or antibiotics; or substance abuse; and 7) injury to non-dominant hand or presence of open skin lesions, disturbed sensation, carpal tunnel syndrome or rash. Women during pregnancy or within 3 months postpartum period were also excluded. The eligibility criteria of HCs were the same as those for IBS participants except that healthy controls did not have IBS.

### Enrollment and Randomization

If eligible, the candidate was scheduled for a study enrollment visit by one of the two study coordinators to obtain informed consent and baseline measures. Written consent was obtained from each participant.

A reminder email was sent to the participants 1 to 3 days before each subsequent appointment. Up to three reminders were sent if there was no response from the participants in accordance with the IRB approved study protocol. Participants were considered as a lost contact and/or drop out if no action and/or response was taken after three reminders.

Allocation of the eligible IBS participants to each intervention group (SMOM vs. NL + SMOM) was completed using a stratified and blocked randomization scheme. First, to obtain an approximately equal ratio of female to male in each group, sex was considered a stratification factor by generating a separate block for each sex. Then using a block size of 4, the eligible participants were included either in the SMOM group or the NL+SMOM group. For this purpose, the random number generated by a statistician was saved in an envelope and was drawn by an unblinded study coordinator who was responsible for scheduling participants and administering the study interventions. All the other study team members were blinded to the group allocation, and dummy codes were used to code the dataset to decrease the possibility of determining the randomization scheme. The full description of randomization and blinding was previously described in the published study protocol (Cong, 2018).

### Interventions

#### SMOM

The SMOM intervention was developed by the research team, which consisted of 10 videos with content including IBS-related pain neurophysiology and the brain-gut axis, triggers of IBS-related pain and IBS pain SM strategies (progressive muscle relaxation, guided imagery, mindfulness, belly breathing, pain problem-solving), and advice to increase physical activity (Cong, Ramesh, et al., 2018). Each video lasts around 15 minutes. The SMOM was sent to all the IBS participants subsequently after the enrollment and baseline (T0) data collection.

Once the participants accepted the invitation of the first video, each of the following 9 videos were automatically sent to the participants on subsequent days. Up to three reminders were sent to the participants if they did not watch the video on time. The video links of the SMOM and the reminders were delivered through Research Electronic Data Capture (REDCap). The links of the videos were also available to the participants at the end of the second week to help them access these materials in case they wanted to review the self-management strategies.

#### NL + SMOM

The NL+SMOM intervention included three sessions of nurse-led one-to-one consultation in addition to the SMOM. Three registered nurses conducted and managed these consultation sessions. Before the study began, the three registered nurses received trainings through mock interviews to keep the consistence of the delivery of the consultation. Each consultation was delivered by a scheduled phone call and lasted around 20 to 30 minutes.

Consultation fidelity was assessed by completing the consultation checklist during each phone call. The first consultation was conducted at the end of the second week once a participant watched the 10 SMOM videos. The second and third consultations were scheduled after the 6- and 12-week follow-up data collection. The first and second nurse-led consultations guided the participants to create their self-management goals (using the SMART format, Supplementary file 1) and to solve any challenges or barriers and modify the self-management goals once reached (Supplementary file 1, consultation guideline). The third consultation debriefed on the accomplishments of the participants at the end of the study. A daily dairy link was also sent to the participants in the NL + SMOM group through the REDCap, by which the participants narratively recorded their IBS related pain, stress, sleep, daily activities, food intake, and stool patterns using Bristol stool scale (Lewis & Heaton, 1997).

### Measurements

Measurements of the current study included demographic characteristics, pain, symptoms, quality of life, self-efficacy, and coping. The demographic characteristics include age, sex, ethnicity, race, education level, and other factors that were measured by the National Institute of Nursing Research (NINR) common data elements (CDEs) (Page et al., 2018).

**Average pain intensity and pain interference** were measured by using the Brief Pain Inventory (BPI). The BPI has questions with 0-10 rating scales, which a higher score refers to more suffer from pain (Keller et al., 2004).

**IBS Quality of life (QOL)** was measured by a 34-item IBS specific QOL instrument (Hahn et al., 1997). The IBS-QOL was designed to capture patients’ perception of their daily functions interfered by IBS. There are 8 subscales in this five-point Linkert instrument. The score range of IBS-QOL is 0-100, which higher score refers to a better QOL.

**IBS related Symptoms** including anxiety, depression, fatigue, and sleep disturbance were measured by using the NIH Patient-Reported Outcomes Measurement Information System (PROMIS®) and scored following the system instruction. A higher T score PROMIS measurement indicated a higher intensity of the measured symptom, and a mean score greater than 55 indicates that a study subject experienced significant higher intensity of the symptom than those of the healthy reference population according to the PROMIS guide (Cella et al., 2010).

**Self-efficacy** was measured by the 6-item Self-Efficacy for Managing Chronic Disease (SEMCD) (Lorig et al., 2001) The score of the Likert SEMCD ranged from 0 to 10. A higher score of SEMCD indicates great self-efficacy.

**Coping strategies** were evaluated by the Coping Strategies Questionnaire-Revised (CSQ-R) to assess six cognitive coping strategies to pain, including distraction, catastrophizing, ignoring pain sensations, distancing from pain, coping self-statements, and praying (Robinson et al., 1997). Each subscale has a score range of 0-36, with higher score indicating a better coping strategy.

### Data collection

Data were collected at the research laboratory. Questionnaires were completed online through the REDCap system using a laptop or iPad. The IBS participants filled all the questionnaires at enrollment (T0), 6- and 12-week follow-up visits (T1 and T2), respectively. The HCs completed surveys regarding demographic information, BPI, and PROMIS measurements at enrollment (T0) and at the 12-week follow-up visit (T2).

### Data analysis

Data analyses were performed using the R software (version 4.1.0). Demographic and clinical characteristics were summarized using descriptive statistics for the NL+SMOM, SMOM, and HC groups. The first hypothesis was carried out by testing the difference of outcomes between the baseline and the 12-week visits for the NL+SMOM and the SMOM groups, respectively. A linear mixed model (LMM) was developed for each outcome with subject-level random intercept using the R package “lme4” (Bates et al., 2007). We included group, visit, and the group-by-visit interaction term as the covariates, and added age, sex, race, ethnicity, employment, and year of IBS diagnosis in the models to control the potential confounding effects. For each model, specific testing contrast was constructed for each intervention group.

The second hypothesis was analyzed by testing the significance of the interaction term between group and the 12-week visit in each LMM. This interaction term directly estimated the difference of intervention effect at the 12-week follow up. Furthermore, we implemented longitudinal mediation analyses using the R package “mediation” (Tingley et al., 2014) to explore if coping catastrophizing and self-efficacy mediated the intervention effect on pain and QOL to address the third hypothesis. Statistical inferences of the indirect effects were conducted by constructing 95% confidence interval based on 5,000 bootstrap samples. The mediators, “Self-Efficacy for Managing Chronic Disease” and “coping-catastrophizing” were selected based on the preliminary baseline mediation analysis (Chen et al., 2022).

## Results

### Recruitment and retention

Figure 1 shows the CONSORT flow diagram of the IBS subjects in this study. Among 112 screened, 96 met the inclusion criteria, 80 were recruited, 41 were randomly assigned in the SMOM group, 39 were in the NL + SMOM group, and 80 finished the baseline (T0) data collection. Thirty-five and 27 participants completed data collection at the T1 follow-up visit in the SMOM and NL + SMOM, respectively. Fifty-six completed the T2 follow-up visit with 30 in the SMOM and 26 in the NL + SMOM. The overall retention rate at the 12-week of this study was 70.00% (66.67% in the NL + SMOM and 73.17% in the SMOM).

**Figure 1.**
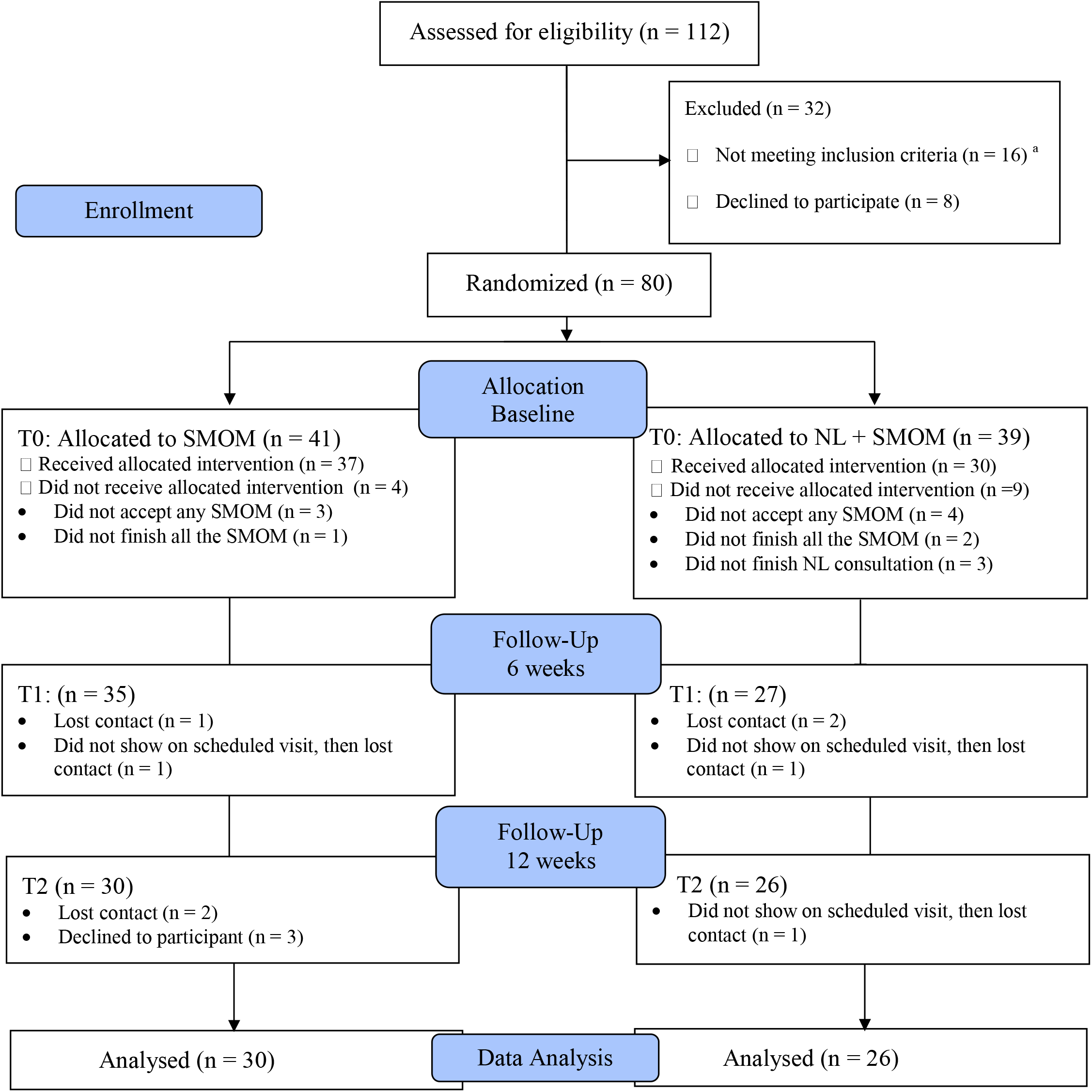
CONSORT flow chart of the current study. SMOM, Self-Management Online education and learning Modules; NL, Nurse-Led. ^a^ Sixteen were ineligible, due to the following reasons: 7 had inflammatory bowel disease/ celiac disease; 2 had chronic pain in another area; 1 had chronic pain in another area and antibiotic use; 1 had chronic pain in another area and other medical conditions (GERD); 2 older than 29 years old; 1 older than 29 years old and other medical conditions (diabetes, bipolar); 1 did not have IBS diagnosis from provider; 1 had other medical conditions (anxiety, asthma, heart problems).

For the HC group recruitment and follow up, 27 were screened and 21 who met the inclusion criteria were recruited. All 21 recruited participants completed the baseline session. After the baseline assessment, four participants dropped out for the 12-week follow up session and 17 (80.95%) participants completed the final session.

### Participant characteristics

The demographic characteristics are summarized in Table 1. Most IBS subjects were non-Hispanic/Latino White, female, and students who received college or associate degree education. The mean age of IBS subjects and HCs was 21 and 20.14 years old, respectively. There was no significant difference between the two IBS groups in demographic characteristics.

**Table 1.** Demographic characteristics.

|  | NL + SMOM (n = 39) |  | SMOM (n = 41) |  | HC (N = 21) |  |
| --- | --- | --- | --- | --- | --- | --- |
|  | N | Percentage (%) | N | Percentage (%) | N | Percentage (%) |
| Sex |  |  |  |  |  |  |
| Female | 32 | 82.05 | 29 | 70.73 | 11 | 52.38 |
| Male | 7 | 17.95 | 12 | 29.27 | 10 | 47.62 |
| Race |  |  |  |  |  | * |
| White | 31 | 79.49 | 31 | 75.61 | 9 | 42.86 |
| Asian | 3 | 7.69 | 7 | 17.07 | 6 | 28.57 |
| Black or African-American | 5 | 12.82 | 3 | 7.32 | 4 | 19.05 |
| Not reported | 0 | 0 | 0 | 0 | 2 | 9.52 |
| Ethnicity |  |  |  |  |  |  |
| Not Hispanic or Latino | 34 | 87.18 | 34 | 82.93 | 16 | 76.19 |
| Hispanic or Latino | 3 | 7.69 | 4 | 9.76 | 4 | 19.05 |
| Not reported | 2 | 5.13 | 3 | 7.32 | 1 | 4.76 |
| Education |  |  |  |  |  |  |
| High school or below | 2 | 5.13 | 4 | 9.76 | 2 | 9.52 |
| College or associate degree | 26 | 66.66 | 22 | 55.36 | 17 | 80.95 |
| Bachelor degree | 7 | 17.95 | 7 | 17.07 | 2 | 9.52 |
| Graduate or Doctorate degree | 4 | 10.26 | 8 | 19.51 | 0 | 0 |
| Employment Status |  |  |  |  |  |  |
| Student | 27 | 69.23 | 30 | 73.17 | 18 | 85.71 |
| Working now | 10 | 25.64 | 10 | 24.39 | 2 | 9.52 |
| Unemployed or other | 2 | 5.13 | 1 | 2.44 | 1 | 4.76 |
| Marital Status |  |  |  |  |  |  |
| Never married | 38 | 97.44 | 39 | 4.88 | 21 | 100 |
| Married | 1 | 2.56 | 2 | 95.12 | 0 | 0 |
|  | Mean | SD | Mean | SD | Mean | SD |
| Age | 21.23 | 2.40 | 21.54 | 2.75 | 20.14 | 1.39 |
| Year of IBS diagnosis | 3.03 | 2.86 | 3.20 | 3.20 |  |  |

NL, Nurse-Lead; SMOM, Self-Management Online education and learning Modules; HC, healthy control.
There was no significant difference regarding the demographic characteristics between two IBS groups.

There were less non-Hispanic White participants in the HC group compared with the IBS groups (p = 0.018).

The longitudinal trends of pain and symptom measurements in the three groups are displayed in Figure 2 and Supplementary Table 1. Compared with the HC group, the IBS participants reported significantly higher BPI average pain intensity and pain interference at both baseline and 12-week visits and intensive anxiety, fatigue, and sleep disturbance at baseline. At the 12-week visit, significant decreasing trend was observed in the average pain intensity and pain interference in both the SMOM and NL + SMOM groups, while a decreasing trend of anxiety was apparent only in the NL + SMOM group.

**Figure 2.**
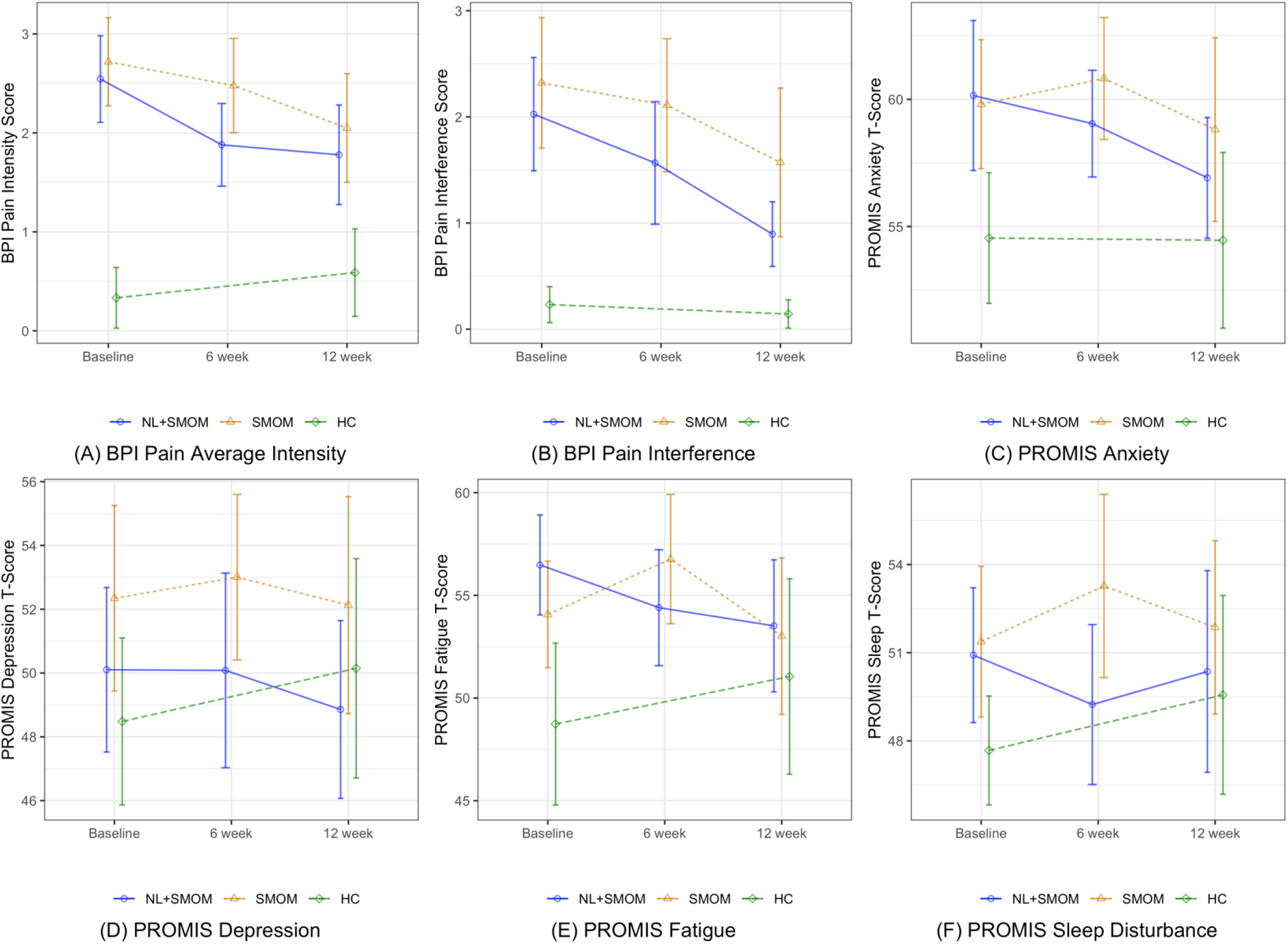
Comparison of pain and symptoms among the three groups. NL, Nurse-Led; SMOM, Self-Management Online education and learning Modules; HC, healthy control. (A) Comparison of BPI average pain intensity between NL + SMOM, SMOM and HC at baseline, 6-week, and 12-week visits, respectively; (B) Comparison of BPI pain interference between NL + SMOM, SMOM and HC at baseline, 6-week, and 12-week visits, respectively. (C) Comparison of PROMIS anxiety T-score between NL + SMOM, SMOM and HC at baseline, 6-week, and 12-week visits, respectively; (D) Comparison of PROMIS depression T-score between NL + SMOM, SMOM and HC at baseline, 6-week, and 12-week visits, respectively; (E) Comparison of PROMIS fatigue T-score between NL + SMOM, SMOM and HC at baseline, 6-week, and 12-week visits, respectively; (F) Comparison of PROMIS sleep disturbance T-score between NL + SMOM, SMOM and HC at baseline, 6-week, and 12-week visits, respectively.

The temporal changes of IBS-QOL levels in the two IBS groups are shown in Supplementary Figure 1 and Supplementary Table 1. Significant increasing trends were observed for the NL + SMOM intervention between baseline and the 12-week in the IBS-QOL total score and the subscales of dysphoria, interference with activity, body image, health worry, social reaction, and relationship. In contrast, the IBS-QOL subscale scores among the SMOM group were not significantly different over time.

### Intervention effects on pain, IBS-QOL, and symptoms

Table 2 displays the estimates of intervention effects on pain, QOL, and symptom measurements at the 12-week. The NL + SMOM intervention significantly decreased the average pain intensity (b = -0.730, p = 0.003), pain interference (b = -1.194, p < 0.001), anxiety score (b = -3.433, p = 0.016), and increased the IBS-QOL score (b = 10.49, p < 0.001). The NL + SMOM intervention also slightly reduced the fatigue score (b = -2.238, p = 0.098) of the IBS participants. For the SMOM only group, the intervention significantly reduced IBS-related pain for average pain intensity (b = -0.592, p = 0.009) and pain interference (b = -0.770, p = 0.008), and improved the IBS-QOL score (b = 4.364, p = 0.033) as well. However, the SMOM intervention did not show significant improvement on any symptom outcomes. No adverse events were reported from participants in either group.

**Table 2.** Linear mixed model results for the comparison of outcome variables between the two IBS groups.

| Group | Intervention effect at 6 weeks<br>(Reference: Baseline) |  | Intervention effect at 12 weeks<br>(Reference: Baseline) |  | Group difference of intervention effect<br>at 12 weeks (reference: SMOM) |  |
| --- | --- | --- | --- | --- | --- | --- |
|  | Estimate (b) | p-value | Estimate (b) | p-value | Estimate (b) | p-value |
| **BPI Average Pain Intensity** |  |  |  |  |  |  |
| NL + SMOM | -0.638 | 0.007 | -0.730 | 0.003 | -0.138 | 0.674 |
| SMOM | -0.209 | 0.331 | -0.592 | 0.009 |  |  |
| **BPI Pain Interference** |  |  |  |  |  |  |
| NL + SMOM | -0.428 | 0.153 | -1.194 | <0.001 | -0.424 | 0.311 |
| SMOM | -0.259 | 0.344 | -0.770 | 0.008 |  |  |
| **IBS-QOL: Total Score** |  |  |  |  |  |  |
| NL + SMOM | 8.031 | <0.001 | 10.49 | <0.001 | 6.125 | 0.040 |
| SMOM | 2.791 | 0.151 | 4.364 | 0.033 |  |  |
| **PROMIS Anxiety T-score** |  |  |  |  |  |  |
| NL + SMOM | -1.236 | 0.368 | -3.433 | 0.016 | -3.435 | 0.086 |
| SMOM | 1.509 | 0.232 | -0.088 | 0.932 |  |  |
| **PROMIS Depression T-score** |  |  |  |  |  |  |
| NL + SMOM | -0.341 | 0.782 | -1.883 | 0.137 | -1.823 | 0.286 |
| SMOM | 0.594 | 0.590 | -0.060 | 0.956 |  |  |
| **PROMIS Fatigue T-score** |  |  |  |  |  |  |
| NL + SMOM | -1.120 | 0.387 | -2.238 | 0.098 | -1.469 | 0.423 |
| SMOM | 2.843 | 0.020 | -0.770 | 0.534 |  |  |
| **PROMIS Sleep Disturbance T-score** |  |  |  |  |  |  |
| NL + SMOM | -1.013 | 0.397 | -1.053 | 0.394 | -0.866 | 0.597 |
| SMOM | 1.367 | 0.209 | -0.187 | 0.883 |  |  |

IBS, irritable bowel syndrome; SMOM, Self-Management Online education and learning Modules; BPI, brief pain inventory; NL, Nurse-Lead; QOL, quality of life.

### Enhanced intervention effect of the nurse-led intervention

Table 2 presents the difference of intervention effects at 12-week between the NL + SMOM and the SMOM groups. There was no significant difference of the positive effect on reducing average pain intensity and pain interference between the two interventions (NL + SMOM vs. SMOM). However, the NL + SMOM significantly improved more IBS-QOL (b = 6.125, p = 0.040) compared with the SMOM intervention alone. The estimated differences of the intervention effects between the two groups on IBS-QOL subscales are shown in Supplemental Table 2. The NL + SMOM enhanced the IBS-QOL with respect to dysphoria (b = 12.977, p = 0.002), health worry (b = 8.842, p = 0.018), and relationship (b = 7.104, p = 0.029). Moreover, the NL + SMOM group slightly reduced anxiety (b = -3.435, p = 0.086) compared with the SMOM group (Table 2).

### Self-management and the mechanisms of the interventions

The self-management strategy measurements of the two IBS groups in the three visits are displayed in Supplementary Table 3. The NL + SMOM group demonstrated an increasing trend in self-efficacy measured by the SEMCD and a decreasing coping praying score at 12-week, while the SMOM group reported a decreasing coping catastrophizing score at the 12-week visit.

The longitudinal mediation analysis in Figure 3 shows that the effects of these two interventions on reducing pain and improving IBS-QOL were mediated by different self-management mechanisms. Self-efficacy was a significant mediator of the NL + SMOM intervention effect, while coping catastrophizing was a significant mediator of the SMOM intervention effect. Specifically, self-efficacy mediated 19.21% (indirect effect = -0.225, 95% CI = [-0.447, -0.033]) of the NL + SMOM intervention effect on reducing pain interference, and mediated 19.19% (indirect effect = 2.158, 95% CI = [0.298, 4.424]) of the NL + SMOM intervention effect on improving IBS-QOL. In contrast, the decrease in coping catastrophizing mediated 55.85% (indirect effect = -0.377, 95% CI = [-0.707, -0.065]) of the SMOM intervention effect on reducing pain interference and mediated 80.61% (indirect effect = 3.293, 95% CI = [0.514, 6.352]) of the SMOM intervention effect on improving IBS-QOL.

**Figure 3.**
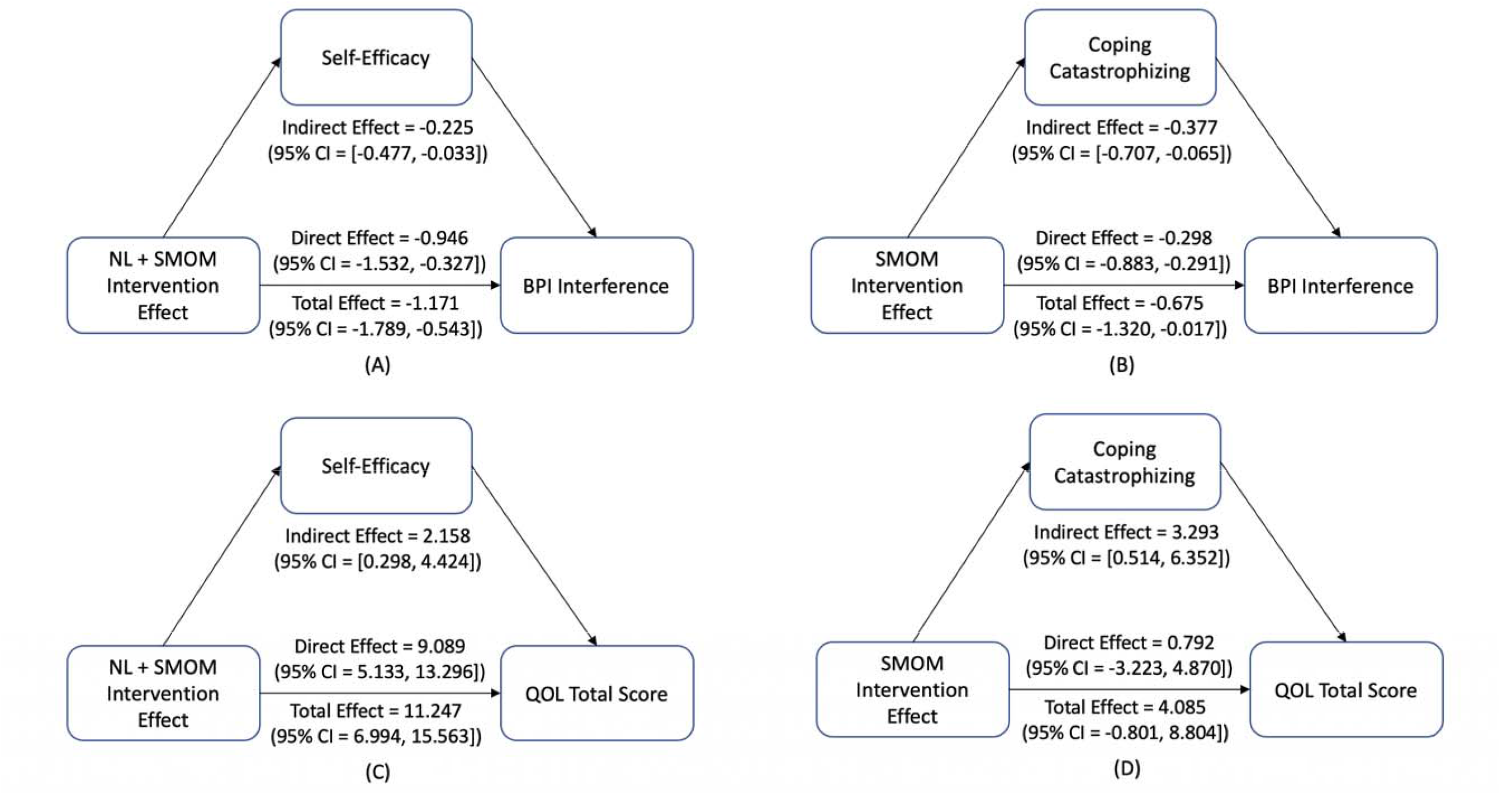
Direct and indirect effect of interventions on pain and QOL in young adults with IBS by mediation analysis. NL, Nurse-Led; SMOM, Self-Management Online education and learning Modules; QOL, quality of life. (A) Direct and Indirect Effect of NL + SMOM on pain interference; (B) Direct and Indirect Effect of SMOM on pain interference; (C) Direct and Indirect Effect of NL + SMOM on QOL; (D) Direct and Indirect Effect of SMOM on QOL.

## Discussion

By applying the IFSMT as a theoretic framework, the current RCT tested the effect of a nurse-led one-to-one consultation plus self-management education among young adults with IBS. All the IBS participants in this study received an intervention, SMOM or NL + SMOM. Both IBS groups reported significant pain relief and IBS-QOL improvement at the 12-week follow up compared with the baseline measurement, partially supporting the primary hypothesis.

Having a diagnosis of IBS at a young age can be overwhelming especially when left to navigate symptom management on their own (Tosic-Golubovic et al., 2010). Lack of self-management knowledge and skills has been a challenge for young adults with IBS (Enck et al., 2016; Hollier et al., 2018). The SMOM developed by our team had been established to be an effective approach of self-management among young adults with IBS. The SMOM group reported significantly decreased pain intensity and interference (Table 2, Fig 2 A and 2 B) after viewing the modules. The results reflect that the 10 online modules including approximately 15 minutes of content each are appropriate for this group of young adults, e.g., delivering via internet, in easy-to-follow format, and the short duration allowing them to hold interest (Pedersen, 2015).

Participants in the NL + SMOM group also reported better management of pain and anxiety, and IBS-QOL improvement at follow up visits compared with the SMOM alone group. Thus, the results supported our second hypothesis that NL + SMOM would have a significantly greater interventional effect on managing pain and enhancing QOL comparing with SMOM only. The nurse-led one-to-one consultation in this RCT not only focused on pain self-management, but also addressed stress management, as well as self-management goal setting. Pain self-management strategies were reflected and reinforced during the nurse-led one-to-one consultations. The consultations guided participants in setting their pain self-management goals, which appeared to be an effective way to help participants activate their newly acquired self-management knowledge and skills. Participants in the NL + SMOM group have a quicker benefit than the SMOM only group since the subjects in NL + SMOM reported improved pain management and QOL at 6-week follow up, while the SMOM group did not at the same time frame. The findings were consistent with a previous study which reported nurse-led self-management enhanced pain and symptom management among patients with IBS through education, coaching and consulting (Niesen et al., 2018). Further studies could compare the cost-effectiveness of nurse-led self-management interventions among IBS subjects and/or evaluate the impact on self-management actions, healthcare visits, and utilization of other support services.

Among the IBS related symptoms, the mean score of anxiety was higher than 55 at the baseline visit of IBS participants (Supplementary Table 1), which indicated that they had a higher level of anxiety than the healthy reference population (Cella et al., 2010). Other symptom measures (depression, fatigue, and sleep disturbance) were similar to the healthy reference population, e.g., lower than or around 55 at all the visits (Supplementary Table 1). Our results showed that the anxiety level in the NL + SMOM intervention group decreased at the 12-week follow up compared with baseline, indicating the effect of nurse-led consultation session on relieving anxiety. The mechanism of anxiety reduction may be related to the increased self-efficacy, decreased pain intensity and pain interference among subjects in the NL + SMOM group (Shahabi et al., 2016; Ten Brink et al., 2021). Our results also suggest that an intervention targeting a worse symptom (e.g., anxiety in IBS individuals) may yield a detectable interventional effect. Other symptoms (e.g., fatigue and sleep disturbance) that were within normal range according to the reference group may have improved due to the intervention but were not detectable. Other reason could be the absence of modules in sleep hygiene and mood management. Further studies could also develop additional modules to address the unmet needs of symptom management among IBS population.

The results from mediation analysis supported the third hypothesis that the NL + SMOM and SMOM interventions improved pain management and IBS-QOL through modifying coping strategies and self-management of chronic disease. Moreover, our results indicate that the NL + SMOM and SMOM alleviated pain and improved QOL through different indirect mechanisms, i.e., by increasing self-efficacy for the NL + SMOM and decreasing catastrophizing for SMOM alone, respectively (Figure 3). This was consistent with the change of self-efficacy and coping- catastrophizing. The SMOM mediated pain and QOL by decreasing inefficient coping strategies (catastrophizing) which indicates that the SMOM is a reliable resource to help young adults effectively cope with their IBS and improve self-management. Previous studies also reported that higher self-efficacy was associated with less pain and higher QOL, higher utilization of catastrophizing was associated with higher pain and lower QOL (Lorig et al., 2001; Shahabi et al., 2016; Ten Brink et al., 2021) .

Although both IBS intervention groups reported decreased pain at the 12-week follow up visit, their average pain intensity and pain interference were still significantly higher than those measured in the HCs. This improvement in pain and symptom management as well as IBS-QOL does not guarantee that the effect will last longer than 3 months. A previous study demonstrated that the effect of a IBS symptom management intervention faded at 6-month follow up (Shahabi et al., 2016). IBS is a chronic condition which requires life-long self-management (Vasant et al., 2021). The present study demonstrates that nurse-led one-to-one consultation benefited young adults with IBS greater than just receiving the information (i.e., SMOM only group). Future studies could evaluate implementation of the intervention in clinical practice settings and assess outcomes beyond 3 months.

This study tested the effect of two interventions on managing pain underpinning the theoretical framework of IFSMT. Results supported the theoretic hypotheses that self-management intervention had direct and indirect effects on decreasing pain and improving IBS-QOL (Ryan & Sawin, 2009; Shahabi et al., 2016). Further studies could test the effect of our education modules as well as interventions underpinning the IFMST among more racially and ethnically diverse groups of young adults with IBS, or patients in middle age or older adults with IBS.

## Limitation

Several limitations emerged in the current study. Only participants with access to the internet were recruited in this study. IBS subjects without daily internet access may have more urgent needs for pain management. Most of the subjects enrolled in the current study were non-Hispanic White female, which is not reflective of the entire IBS population. Due to the nature of a pilot study, the multiplicity adjustment was not performed in testing multiple hypotheses to control the family-wise type I error. Further confirmatory studies with longer follow-up period could recruit a more diverse population of IBS and engage communities with limit recourses.

## Conclusion

This RCT examined the effect of self-management online modules (SMOM) and a nurse-led one-to-one consultation plus SMOM (NL + SMOM) on pain self-management among young adults with IBS. Both the NL + SMOM and SMOM alone showed significant interventional benefits on pain relief and IBS-QOL improvement at the follow up visits. The NL + SMOM also decreased anxiety among IBS subjects. Further studies could follow up the participants at a longer interval since IBS as a chronic condition that needs life-long self-management. These interventions could also be employed in resource limited settings such as underserved communities to improve population health.

## Data Availability

All data produced in the present study are available upon reasonable request to the authors.

## Author Contributions

Conceptualization, J.C., X.C., Y.Z., W. X., Z. A. B., B. F., and A. S.; formal analysis, Y.Z., J.C., W. X., Z. A. B., J. L., T.Z., M.H.C., and X.C.; funding acquisition, X. C., and A. S.; methodology, J.C., W. X., Z. A. B., J. L., M.H.C., and X.C.; project administration, ., W. X., and X.C.; writing-original draft, J.C., Y.Z., Z. A. B., T.Z., W. X., J. L., and X.C.; writing- review and editing, All. All authors have reviewed the manuscript and agreed to submit this version.

## Acknowledgments

The authors would like to acknowledge all the participants in this study. The authors would also like to acknowledge the support from the Bio-Behavioral Lab (BBL), the Center of Advancement in Managing Pain (CAMP), and NIH funded P20 Center for Accelerating Precision Pain Self-Management in the University of Connecticut School of Nursing.

---

**Supplementary Figure 1.**
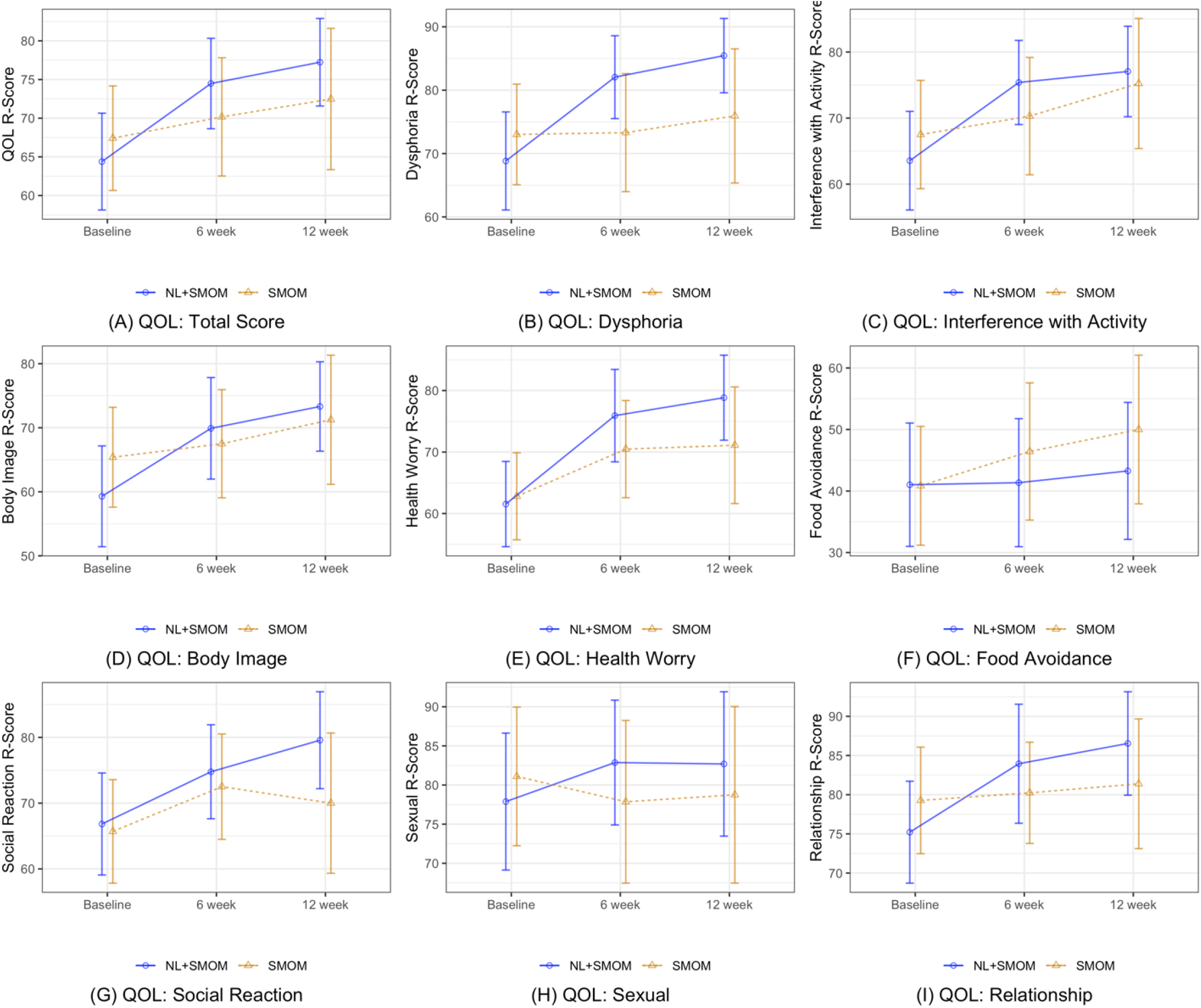
Longitudinal trends of IBS-QOL total score and subscales of the two IBS groups. NL, Nurse-Led; SMOM, Self-Management Online education and learning Modules; QOL, quality of life. (A) Comparison of QOL total score between NL + SMOM and SMOM at baseline, 6 weeks, and 12 weeks visits, respectively; (B) Comparison of QOL Dysphoria sub-scale score between NL + SMOM and SMOM at baseline, 6 weeks, and 12 weeks visits, respectively; (C) Comparison of QOL Interference with Activity sub-scale score between NL + SMOM and SMOM at baseline, 6 weeks, and 12 weeks visits, respectively; (D) Comparison of QOL Body Image sub-scale score between NL + SMOM and SMOM at baseline, 6 weeks, and 12 weeks visits, respectively; (E) Comparison of QOL Health Worry sub-scale score between NL + SMOM and SMOM at baseline, 6 weeks, and 12 weeks visits, respectively; (F) Comparison of QOL Food Avoidance sub-scale score between NL + SMOM and SMOM at baseline, 6 weeks, and 12 weeks visits, respectively; (G) Comparison of QOL Social Reaction sub-scale score between NL + SMOM and SMOM at baseline, 6 weeks, and 12 weeks visits, respectively; (H) Comparison of QOL Sexual sub-scale score between NL + SMOM and SMOM at baseline, 6 weeks, and 12 weeks visits, respectively; (I) Comparison of QOL Relationship sub-scale score between NL + SMOM and SMOM at baseline, 6 weeks, and 12 weeks visits, respectively;

**Supplementary Table 1.**
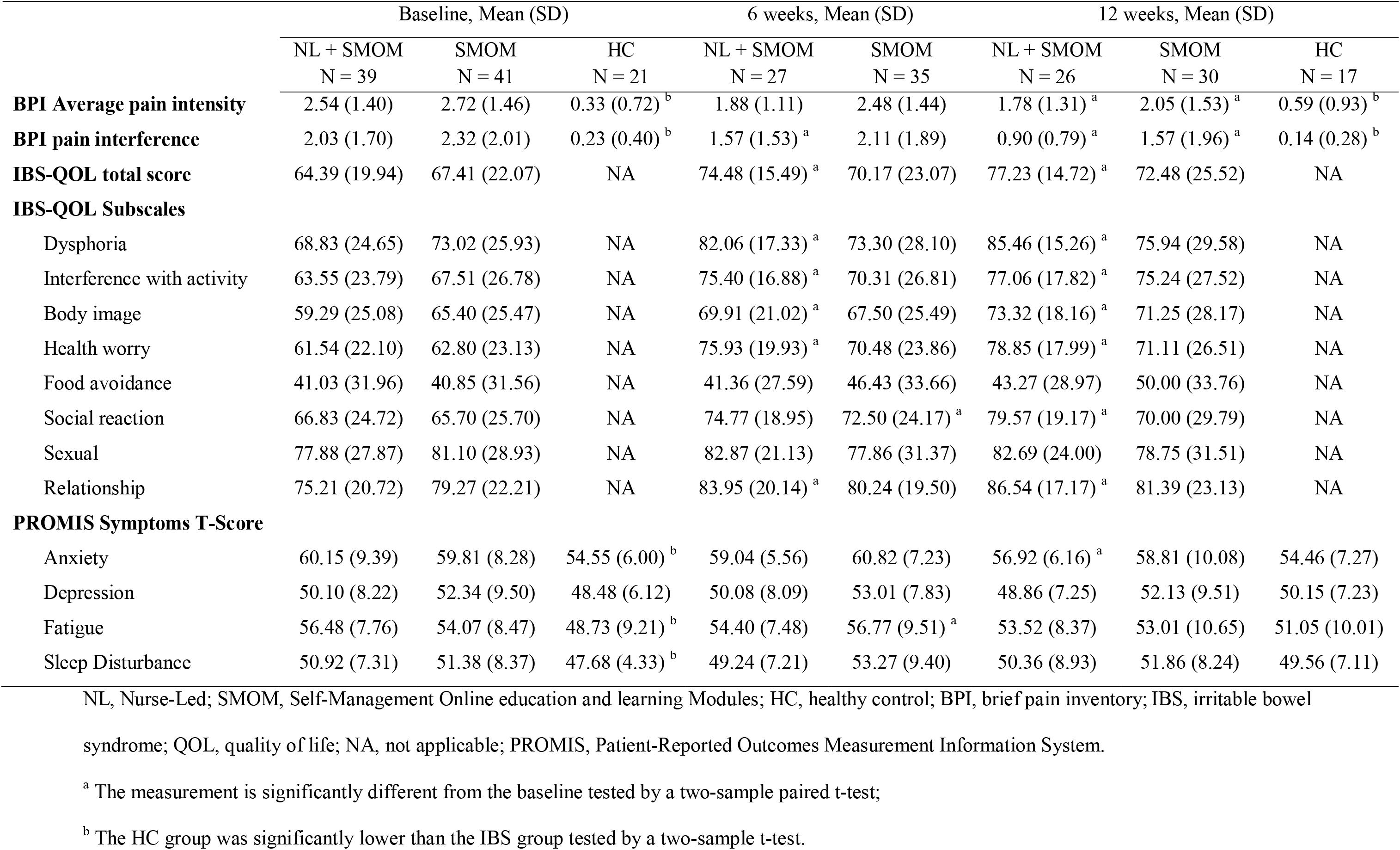
Summary statistics of the outcome variables among the three groups at the three time points.

|  | Baseline, Mean (SD) |  |  | 6 weeks, Mean (SD) |  | 12 weeks, Mean (SD) |  |  |
| --- | --- | --- | --- | --- | --- | --- | --- | --- |
|  | NL + SMOM<br>N = 39 | SMOM<br>N = 41 | HC<br>N = 21 | NL + SMOM<br>N = 27 | SMOM<br>N = 35 | NL + SMOM<br>N = 26 | SMOM<br>N = 30 | HC<br>N = 17 |
| **BPI Average pain intensity** | 2.54 (1.40) | 2.72 (1.46) | 0.33 (0.72)<sup>b</sup> | 1.88 (1.11) | 2.48 (1.44) | 1.78 (1.31)<sup>a</sup> | 2.05 (1.53)<sup>a</sup> | 0.59 (0.93)<sup>b</sup> |
| **BPI pain interference** | 2.03 (1.70) | 2.32 (2.01) | 0.23 (0.40)<sup>b</sup> | 1.57 (1.53)<sup>a</sup> | 2.11 (1.89) | 0.90 (0.79)<sup>a</sup> | 1.57 (1.96)<sup>a</sup> | 0.14 (0.28)<sup>b</sup> |
| **IBS-QOL total score** | 64.39 (19.94) | 67.41 (22.07) | NA | 74.48 (15.49)<sup>a</sup> | 70.17 (23.07) | 77.23 (14.72)<sup>a</sup> | 72.48 (25.52) | NA |
| **IBS-QOL Subscales** |  |  |  |  |  |  |  |  |
| Dysphoria | 68.83 (24.65) | 73.02 (25.93) | NA | 82.06 (17.33)<sup>a</sup> | 73.30 (28.10) | 85.46 (15.26)<sup>a</sup> | 75.94 (29.58) | NA |
| Interference with activity | 63.55 (23.79) | 67.51 (26.78) | NA | 75.40 (16.88)<sup>a</sup> | 70.31 (26.81) | 77.06 (17.82)<sup>a</sup> | 75.24 (27.52) | NA |
| Body image | 59.29 (25.08) | 65.40 (25.47) | NA | 69.91 (21.02)<sup>a</sup> | 67.50 (25.49) | 73.32 (18.16)<sup>a</sup> | 71.25 (28.17) | NA |
| Health worry | 61.54 (22.10) | 62.80 (23.13) | NA | 75.93 (19.93)<sup>a</sup> | 70.48 (23.86) | 78.85 (17.99)<sup>a</sup> | 71.11 (26.51) | NA |
| Food avoidance | 41.03 (31.96) | 40.85 (31.56) | NA | 41.36 (27.59) | 46.43 (33.66) | 43.27 (28.97) | 50.00 (33.76) | NA |
| Social reaction | 66.83 (24.72) | 65.70 (25.70) | NA | 74.77 (18.95) | 72.50 (24.17)<sup>a</sup> | 79.57 (19.17)<sup>a</sup> | 70.00 (29.79) | NA |
| Sexual | 77.88 (27.87) | 81.10 (28.93) | NA | 82.87 (21.13) | 77.86 (31.37) | 82.69 (24.00) | 78.75 (31.51) | NA |
| Relationship | 75.21 (20.72) | 79.27 (22.21) | NA | 83.95 (20.14)<sup>a</sup> | 80.24 (19.50) | 86.54 (17.17)<sup>a</sup> | 81.39 (23.13) | NA |
| **PROMIS Symptoms T-Score** |  |  |  |  |  |  |  |  |
| Anxiety | 60.15 (9.39) | 59.81 (8.28) | 54.55 (6.00)<sup>b</sup> | 59.04 (5.56) | 60.82 (7.23) | 56.92 (6.16)<sup>a</sup> | 58.81 (10.08) | 54.46 (7.27) |
| Depression | 50.10 (8.22) | 52.34 (9.50) | 48.48 (6.12) | 50.08 (8.09) | 53.01 (7.83) | 48.86 (7.25) | 52.13 (9.51) | 50.15 (7.23) |
| Fatigue | 56.48 (7.76) | 54.07 (8.47) | 48.73 (9.21)<sup>b</sup> | 54.40 (7.48) | 56.77 (9.51)<sup>a</sup> | 53.52 (8.37) | 53.01 (10.65) | 51.05 (10.01) |
| Sleep Disturbance | 50.92 (7.31) | 51.38 (8.37) | 47.68 (4.33)<sup>b</sup> | 49.24 (7.21) | 53.27 (9.40) | 50.36 (8.93) | 51.86 (8.24) | 49.56 (7.11) |

NL, Nurse-Led; SMOM, Self-Management Online education and learning Modules; HC, healthy control; BPI, brief pain inventory; IBS, irritable bowel
syndrome; QOL, quality of life; NA, not applicable; PROMIS, Patient-Reported Outcomes Measurement Information System.
<sup>a</sup> The measurement is significantly different from the baseline tested by a two-sample paired t-test;
<sup>b</sup> The HC group was significantly lower than the IBS group tested by a two-sample t-test.

**Supplementary Table 2.**
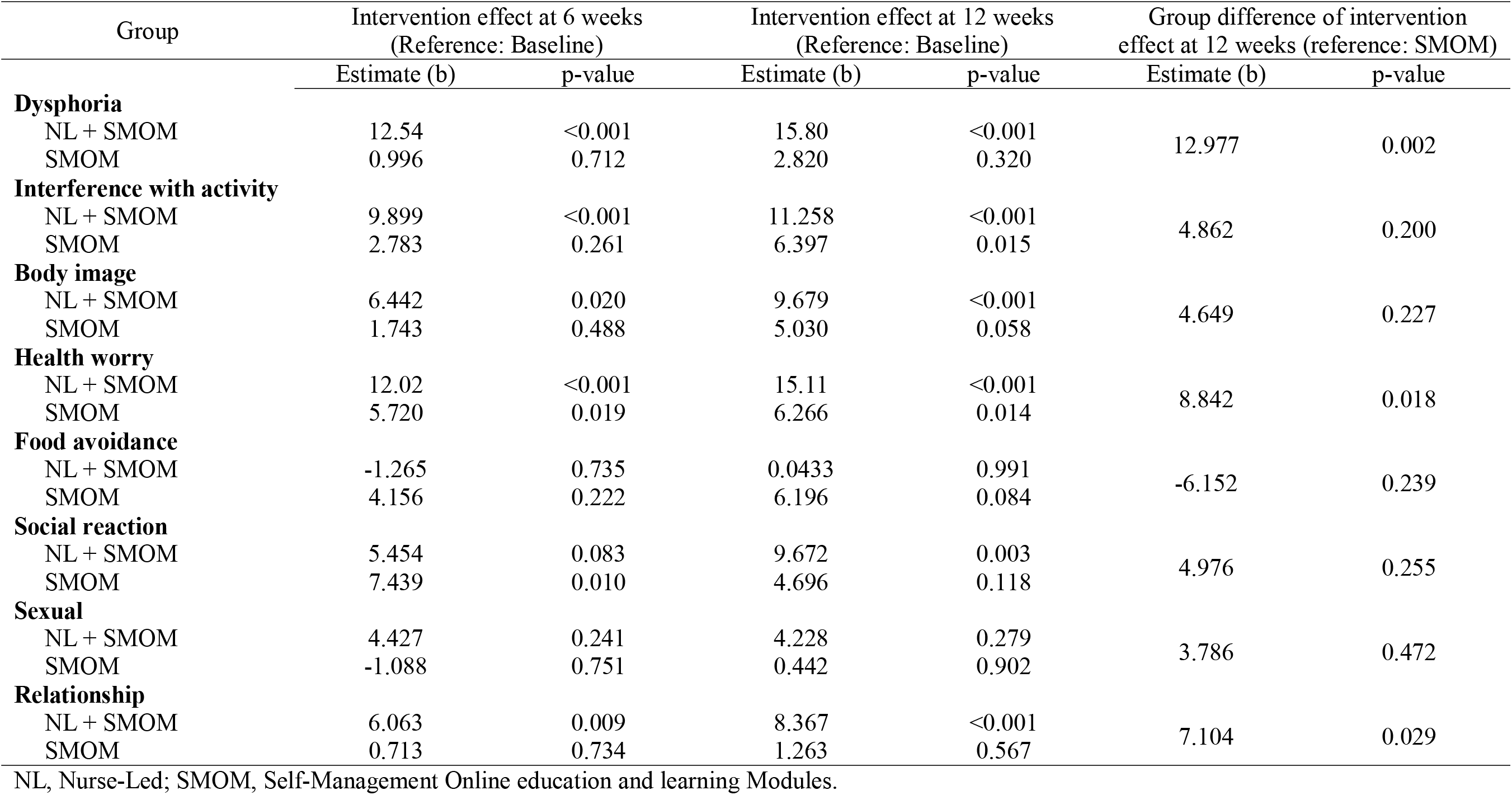
Linear mixed models for IBS-QOL subscales.

**Supplementary Table 3.** Summary statistics of the self-management variables among the IBS subjects at the three time points.

|  | Baseline, Mean (SD) |  | 6 weeks, Mean (SD) |  | 12 weeks, Mean (SD) |  |
| --- | --- | --- | --- | --- | --- | --- |
|  | NL + SMOM | SMOM | NL + SMOM | SMOM | NL + SMOM | SMOM |
| **SEMCD** | 6.66 (1.72) | 6.58 (2.05) | 7.17 (1.69) | 6.50 (2.16) | 7.29 (1.68)<sup>a</sup> | 7.06 (2.17) |
| **Coping Strategy** |  |  |  |  |  |  |
| Self-statement | 4.88 (1.17) | 4.96 (1.30) | 4.69 (1.40) | 4.82 (1.37) | 4.69 (1.40) | 4.82 (1.37) |
| Praying | 2.26 (1.82) | 3.53 (2.19) | 2.10 (1.58) | 2.88 (2.02) | 1.88 (1.41)<sup>a</sup> | 2.73 (2.05) |
| Distancing | 1.88 (1.26) | 2.10 (1.53) | 2.35 (1.32)<sup>a</sup> | 2.29 (1.51) | 2.12 (1.59) | 2.32 (1.42) |
| Ignoring | 3.57 (1.65) | 3.75 (1.49) | 3.67 (1.55) | 4.01 (1.57) | 3.58 (1.64) | 4.15 (1.58) |
| Catastrophizing | 2.38 (1.19) | 2.82 (1.32) | 2.15 (0.99) | 2.56 (1.43) | 2.10 (0.94) | 2.32 (1.30)<sup>a</sup> |
| Distraction | 3.34 (1.53) | 3.51 (1.58) | 3.54 (1.49) | 3.34 (1.52) | 3.55 (1.50) | 3.35 (1.60) |

NL, Nurse-Led; SMOM, Self-Management Online education and learning Modules; SEMCD, self-efficacy for managing chronic disease.
<sup>a</sup> The measurement is significantly different from the baseline tested by a two-sample paired t-test.

