## Supplementary file 1 for "The effect of self-management online modules plus nurse-led support on pain and quality of life among young adults with irritable bowel syndrome: A randomized controlled trial"

**IBS Study Consultation Call**

1. Retrieve the following information from REDCap and calendar
   a. Name
   b. Time of appointment
   c. Visit number/ type of visit
   d. Group (only experimental group needed to call)
   e. Other relevant info
   f. Review subject’s health information, including IRB symptoms (i.e., pain, diarrhea, constipation) and self-management behaviors (i.e., diet, daily activities, medication use).
2. Scripts for calls (about 20 – 30 mins)
   a. Hello, my name is _____ and I’m a registered nurse and a research assistant working with Dr. Cong on the IBS study. I am calling to follow up and provide consultation about self-management of IRB.
   b. **Symptom episodes**: Could you please tell me any symptoms that you're experiencing recently (last two weeks?), such as abdominal pain, bloating, diarrhea, or constipation, and for how server, how long and how often? Does anything seem to trigger your symptoms, including certain foods, stress or in women - your menstrual period? Do you have other health conditions?
   c. **Stress management:** Could you please tell me if there any recent changes or stressors in your life, such as family changes, pressures from school and friends, emotional difficulties, and other issues? (These factors can play a key role in the frequency and severity of IBS symptoms.) How do you cope with the stressors in your life?

**Provide consultation** – relaxation, meditation, stress management

d. **Diet:** What is your typical daily diet? What type of diet do you have recently? Do you have any dietary changes recently? If yes, what kind of change? Does it help to reduce your symptoms?

**Provide education**: high fiber diet, avoiding triggers…

e. **Daily activities:** Could you please tell me what typical daily activities do you enjoy? How many hours of study/work in classroom, lab, or other places? How many minutes for excises? What types of excise (gym, swimming, jogging, hiking…)? Are there any changes of daily activities recently? Does the change to help reduce or manage your symptoms?

**Provide consultation:** 150 minutes/week of moderate to vigorous exercise

f. **Medications**: Could you please tell me what medications you are using now (including prescription and over-the-counter medications, vitamins, herbs, and supplements)? In what condition do you use medications (name)? Have you adhered to the medication regimen? If not, reasons?

**Provide consultation**:

g. **Quality of Life**: Could you please tell me how much would you say your symptoms are affecting your quality of life, such as your personal relationships and your ability to function at school or work? Are you unable to do things that you really enjoy doing? Do you have any concerns?

**Provide consultation**: What are some ways that you can think of to get back into those activities?

h. **Goal Setting:**
   i. What is one way you want to better manage your IBS symptoms and your health?
      1. What is your timeline to focus on this? The next week? The next month?
      2. When will you do this activity? (i.e. Each day in the morning)
      3. What might get in the way from you completing this goal? What will you do if this does stop you?
      4. How successful do you expect to be on this plan? (percentage)
      5. If less than 80%, what can you do to be more successful?
i. **Summary:** Thank you for talking with me. Do you have any additional questions or concerns?
j. **Daily Diary:** *Introduce the daily diary. Link in with baseline questionnaire answers.*
k. **Reminder for next visit and stool sample visit**:

Follow-up visits scheduled for week 6 and week 12 from when you brought in your initial stool sample. They will include taking blood and saliva samples as well as completing the questionnaires and QST testing. You will have another consult with a nurse at both of these visits. We will also collect additional stool samples after those visits. You will be compensated $25 gift card for each visit.

Your next visit will be on __________ (Date/Time)

Please send your stool sample to room XXX in the School of Nursing anytime between the hours of (coverage times) on________ (Date/Time).

Please call at 860-486-XXXX if you have any questions.
